# Universal maternal and child health coverage and its socio-demographic determinants: Findings from the Demographic Health Survey, 2015-16, Afghanistan

**DOI:** 10.64898/2026.07.28.26359184

**Authors:** Khyber Mashal, Norbert Schmitz, Shantanu Sharma

## Abstract

The present study aimed to assess 8 key Maternal and Child Health (MCH) indicators and their socio-demographic determinants in Afghanistan using the 2015 Demographic and Health Survey. It was quantitative analysis on cross-sectional data of ever-married women aged 15-49. The MCH indicators included possession of a health card, birth weight of the child, postnatal check-up, full immunization, exclusive breastfeeding, tetanus toxoid, place of delivery, and antenatal check-up (ANC). We employed multiple correspondence analysis (MCA) and a general linear model to identify associations of MCH indicators with their determinants. The analysis was conducted among 30,000 women. MCA yielded one strong latent construct that accounted for 76.7% of the total variation. The ANC visits, place of delivery, possession of health cards, postnatal visits, and tetanus toxoid vaccination were strongly discriminating items for this dimension. Women who were younger, had a girl child, belonged to the lower wealth quintile, were illiterate, and did not have access to improved drinking water had a higher probability of poor maternal and child health indicators (p<0.05). The MCH situation and their socio-demographic determinants need to be addressed as a priority with concurrent investment in health infrastructure, women’s education, and health literacy of populations.

## Introduction

The maternal mortality ratio (MMR) has decreased drastically in the past two decades, from 339 per 100,000 live births in 2000 to 223 in 2020. Post-2016, the decline in MMR is mainly concentrated in Central and South Asia, Australia, and New Zealand. There has been stagnancy in MMR decline in the east and Southeast Asia and North Africa, which, in fact, contribute significantly to the global MMR prevalence [1]. The global stagnation in MMR is critical to address, as we may fall short of Sustainable Development Goals 2030, and it reflects inequality between and within countries and a flagrant human rights violation [2].

Investments are needed to address the biomedical causes of maternal deaths, implement effective interventions to mitigate the adverse effects of social factors on maternal deaths, and provide universal health coverage. Universal maternal and child health coverage will help ensure quality care during pregnancy, childbirth, and post-pregnancy care [3]. Universal health coverage (UHC), as defined by the World Health Organization, is access to healthcare without financial hardship for everyone in society [4]. The UHC includes a wide spectrum of services, including promotive, curative, rehabilitative, and preventive [5].

Maternal health refers to the status of women during pregnancy, childbirth, and the postnatal period, encompassing routine antenatal, intrapartum, and postpartum care services [6]. Inequalities have been found in maternal health outcomes and care services, especially in Low- and Middle-Income countries. Economic status, area of residence, maternal education and age, ethnicity, and geography are the most identified factors responsible for inequality in access to maternal health services [7]. Hence, ensuring that women have access to respectful, high-quality maternity care is crucial.

Improving maternal health care utilization can not only reduce MMR and maternal morbidity but also improve child health outcomes [8]. There is emerging evidence that the quality-of-care concept should be expanded beyond access to effective coverage. Introduced in 1978, Tanahashi’s model of health system coverage remains an effective coverage framework for assessing maternal and child health outcomes [9]. The Tanahashi’s model includes five key domains, including accessibility, acceptability, contact or actual use, and quality [10].

Afghanistan, a low-income country in central Asia, has one of the worst maternal and child health outcomes globally. After having suffered from decades of war, instability, limited investments in health, and the Taliban government, Afghanistan lies at the bottom of all the health indicators [11]. However, Afghanistan achieved significant improvements between 2000 and 2020, with drastic reductions in MMR, infant mortality, and under-5 mortality, and an expansion of over 2000 functional health centers across the country [12, 13]. Access to health facilities gained significant momentum, with 87% of the population able to reach facilities within 2 hours [14]. However, there is limited published literature and data on access to and utilization of maternal and child health services in Afghanistan, aside from the Demographic and Health Surveys (DHS) [15].

Given the need and scope for transforming maternal and child health outcomes in Afghanistan, we assessed the coverage of maternal and child health services and the factors that affect their utilization. The present study aimed to assess inequalities in the coverage and utilization of maternal and child healthcare services, as well as, their determinants in Afghanistan.

## Materials and Methods

### Study design

It was a cross-sectional quantitative study based on the DHS data of Afghanistan 2015-16 [15]. The data were collected by the central statistical organization in partnership with the Ministry of Public Health, Afghanistan, from June 15^th^ 2015, to February 23^rd^, 2016.

### Sampling

The DHS employed a two-stage stratified sample design to estimate key indicators at the national level across the urban and rural areas of 34 provinces in Afghanistan. In the first stage, 960 clusters, including 260 urban and 690 rural clusters, were selected, followed by the selection of households using systematic random sampling. Around 27 households were selected per cluster using systematic sampling, such that a total of 25,650 households were surveyed.

### Inclusion criteria

All ever-married women aged 15-49 who were either permanent residents of the selected households or visitors who stayed the night before the survey were eligible for the interview.

### Study tool

The Afghanistan DHS 2015-16 included three main questionnaires: the household questionnaire, the women’s questionnaire, and the men’s questionnaire. These were standard DHS questionnaires, contextualized to Afghanistan’s socio-demographic indicators and population health concerns. In the present study, we focused on the women’s questionnaire that had questions on their socio-demographic characteristics, such as their age, education, area of residence (rural or urban), wealth index, number of children, drinking water facility, toilet facility, and the use of cooking fuel (improved or unimproved). Maternal history included questions on the number of antenatal check-ups (ANC), postnatal check-ups (PNC), place of delivery, and tetanus toxoid vaccination of the last pregnancy. In addition, information about the youngest child’s health, such as their age, sex, immunization and the health card status, breastfeeding status, and birth weight were analyzed. Inequalities were assessed based on rural-urban differences. The results were dichotomized into rural and urban categories based on the place of residence of the study participants.

### Ethical clearance

We used secondary anonymized data from the DHS website for Afghanistan 2015-16 (Central Statistics Organization, Ministry of Public Health and ICF 2026). As per the DHS guidelines, necessary ethical approval and written consent were obtained before collecting data from the study population. We obtained ethical approval from the University of Tuebingen, Germany (011/2025BO2) to use secondary data.

### Statistical analysis

We expressed continuous variables as mean and standard deviation, and categorical variables as frequencies and percentages. We identified eight key maternal and child health indicators: health card, birth weight of the child, postnatal check-up, full immunization, exclusive breastfeeding, tetanus toxoid, place of delivery, and antenatal check-up. We employed multiple correspondence analysis (MCA) as the dimension-reduction technique using these 8 indicators. The MCA yields the percent contribution of the dimensions to the total variation/inertia, and the mass, quality, and percent inertia of each variable to the overall model. In addition, coordinate, squared correlation, and the contribution of each variable to the individual dimensions defined by the MCA analysis were reported. Higher values of the coordinates (corr) and contribution (contr) reflected greater association and contribution to the particular dimension, respectively. Likewise, a squared correlation value of >0.5 suggested a strong representation, a value of 0.2-0.5 a moderate association, and a value of <0.2 a weak correlation. The positive and negative signs of the coordinates reflected the direction of association between the variable and the dimension [16].

The dimensions obtained from the analysis were used as dependent variables in a linear regression model to identify associations with their socio-demographic determinants. We assessed the normality of the standardized residuals using the Shapiro-wilk test and Q-Q plot, heteroscedasticity using the Breusch-Pagan test, and multicollinearity using the variance inflation factor (vif). Due to violations of the assumptions for the ordinary linear regression method, we employed the general linear method of regression with family(gaussian), link(identity), and vce(robust) settings. All the analyses were performed using Stata version 14.0 (StataCorp, College Station, TX, USA). A p-value of <0.05 was considered a statistically significant value.

## Results

The analysis was conducted among 30,000 women of reproductive age. Out of 30,000 women, nearly 6056 were pregnant at the time of the survey. The mean age of women was 30 in both rural and urban areas (**Table 1**). Around 70% of women in rural areas depended on wells or rivers for drinking water, while 47% had access to tap water in urban areas. The use of solid fuels, such as dung cakes, wood, charcoal, etc., was common in rural areas (88%).

**Table 1.** Distribution of the socio-demographic factors among women from the rural and urban areas in Afghanistan.

| Variables | Rural (N=22436) N (%) | Urban (n=7025) N (%) |
| --- | --- | --- |
| **Age of women (years); Median (IQR)** | 30 (24-30) | 30 (24-30) |
| **Age of the youngest child (months); Median (IQR)** | 2 (1-4) | 2 (1-5) |
| Missing | 3494 | 855 |
| **Sex of the youngest child** |  |  |
| Boy | 10539 (52.2) | 3382 (52.8) |
| Girl | 9652 (47.8) | 3025 (47.2) |
| Missing | 2245 | 618 |
| **Wealth index** |  |  |
| Poorest | 5236 (23.3) | 411 (5.9) |
| Poorer | 6428 (28.7) | 328 (4.7) |
| Middle | 5879 (26.2) | 477 (6.8) |
| Richer | 4401 (19.6) | 1852 (26.4) |
| Richest | 492 (2.2) | 3957 (56.3) |
| **Educational status of women** |  |  |
| No education | 20113 (89.6) | 5088 (72.4) |
| Primary | 1219 (5.4) | 759 (10.8) |
| Secondary and Higher | 1104 (4.9) | 1178 (16.8) |
| **Total number of children, Median (IQR)** | 4 (2-6) | 4 (2-6) |
| **Type of toilet facility** |  |  |
| Flush latrine (improved) | 984 (4.5) | 2699 (38.7) |
| Pit latrine (unimproved) | 16855 (76.7) | 4088 (58.7) |
| Open | 4127 (18.8) | 179 (2.6) |
| Missing | 470 | 59 |
| **Drinking water source** |  |  |
| Piped or tap (improved) | 6481 (29.6) | 3226 (46.6) |
| Wells or rivers (unimproved) | 15447 (70.4) | 3692 (53.4) |
| Missing | 508 | 107 |
| **Type of cooking fuel** |  |  |
| Gaseous (improved) | 2717 (12.3) | 4747 (68.4) |
| Solid (unimproved) | 19342 (87.7) | 2193 (31.6) |
| Missing | 377 | 85 |

**Abbreviations:** IQR: Interquartile Range

Around 48% of women in rural and 32% in urban areas did not pay a single ANC visit. Around 60% of women in both rural and urban areas had not received any tetanus injection (**Table 2**). Nearly three-fourths of women had never had any PNC visit in both rural and urban areas.

**Table 2.**
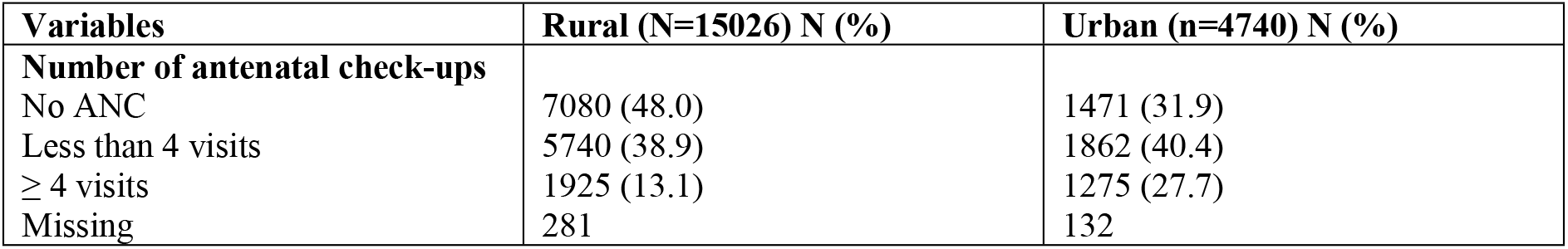
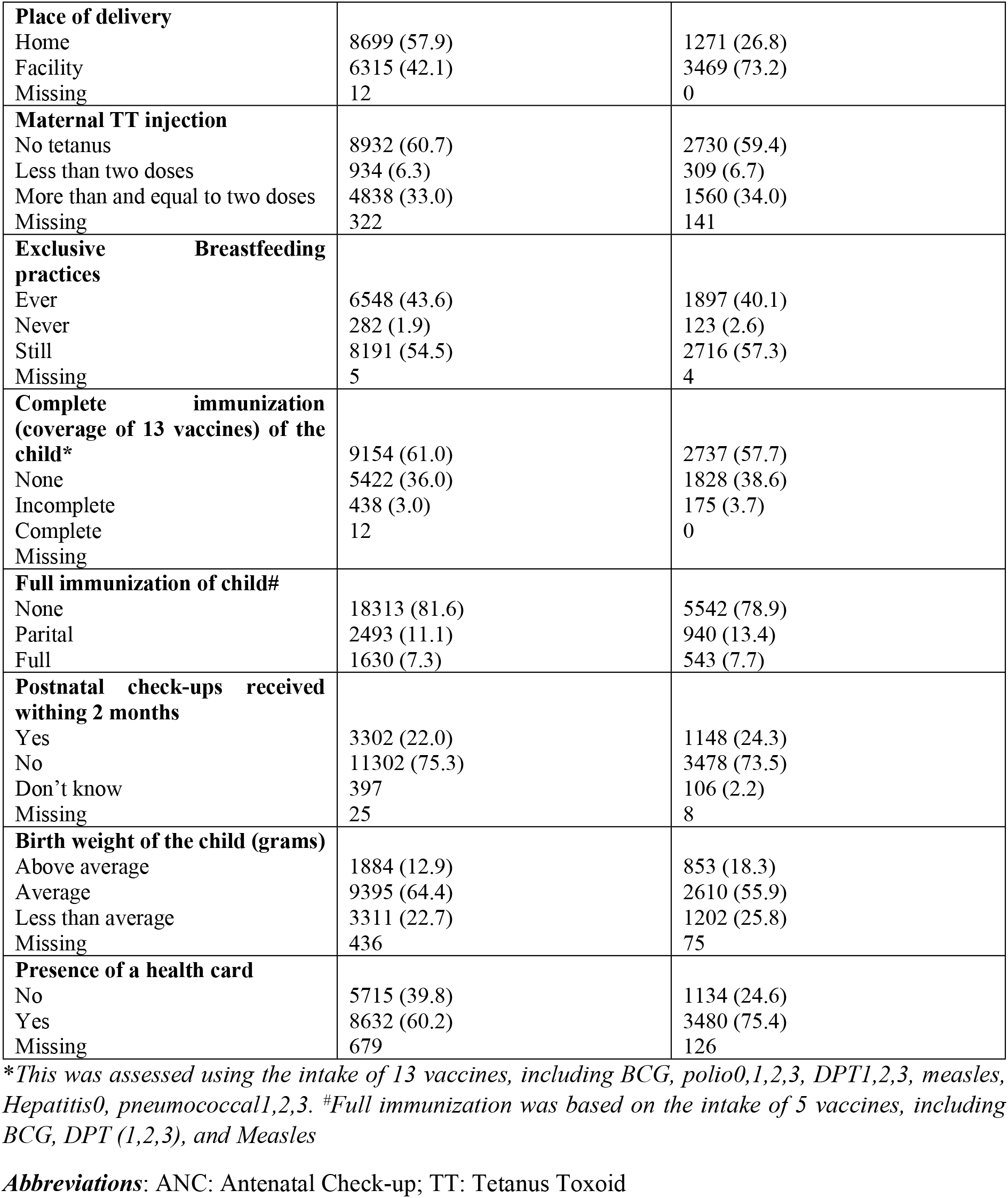
Prevalence of maternal and child health indicators and health services utilization among rural and urban inhabitants in Afghanistan.

| Variables | Rural (N=15026) N (%) | Urban (n=4740) N (%) |
| --- | --- | --- |
| **Number of antenatal check-ups** |  |  |
| No ANC | 7080 (48.0) | 1471 (31.9) |
| Less than 4 visits | 5740 (38.9) | 1862 (40.4) |
| ≥ 4 visits | 1925 (13.1) | 1275 (27.7) |
| Missing | 281 | 132 |
| **Place of delivery** |  |  |
| Home | 8699 (57.9) | 1271 (26.8) |
| Facility | 6315 (42.1) | 3469 (73.2) |
| Missing | 12 | 0 |
| **Maternal TT injection** |  |  |
| No tetanus | 8932 (60.7) | 2730 (59.4) |
| Less than two doses | 934 (6.3) | 309 (6.7) |
| More than and equal to two doses | 4838 (33.0) | 1560 (34.0) |
| Missing | 322 | 141 |
| **Exclusive Breastfeeding practices** |  |  |
| Ever | 6548 (43.6) | 1897 (40.1) |
| Never | 282 (1.9) | 123 (2.6) |
| Still | 8191 (54.5) | 2716 (57.3) |
| Missing | 5 | 4 |
| **Complete immunization (coverage of 13 vaccines) of the child\*** |  |  |
| None | 9154 (61.0) | 2737 (57.7) |
| Incomplete | 5422 (36.0) | 1828 (38.6) |
| Complete | 438 (3.0) | 175 (3.7) |
| Missing | 12 | 0 |
| **Full immunization of child#** |  |  |
| None | 18313 (81.6) | 5542 (78.9) |
| Parital | 2493 (11.1) | 940 (13.4) |
| Full | 1630 (7.3) | 543 (7.7) |
| **Postnatal check-ups received withing 2 months** |  |  |
| Yes | 3302 (22.0) | 1148 (24.3) |
| No | 11302 (75.3) | 3478 (73.5) |
| Don't know | 397 | 106 (2.2) |
| Missing | 25 | 8 |
| **Birth weight of the child (grams)** |  |  |
| Above average | 1884 (12.9) | 853 (18.3) |
| Average | 9395 (64.4) | 2610 (55.9) |
| Less than average | 3311 (22.7) | 1202 (25.8) |
| Missing | 436 | 75 |
| **Presence of a health card** |  |  |
| No | 5715 (39.8) | 1134 (24.6) |
| Yes | 8632 (60.2) | 3480 (75.4) |
| Missing | 679 | 126 |

\*This was assessed using the intake of 13 vaccines, including BCG, polio0,1,2,3, DPT1,2,3, measles, Hepatitis0, pneumococcal1,2,3. #Full immunization was based on the intake of 5 vaccines, including BCG, DPT (1,2,3), and Measles
**Abbreviations:** ANC: Antenatal Check-up; TT: Tetanus Toxoid

The MCA analysis yielded 5 dimensions; however, the first dimension accounted for 76.7% of the total inertia (variations/patterns among 8 categorical variables), while the second accounted for only 4.9%. The other four dimensions accounted for 0.01-0.55% of the total inertia, a weak contribution to the total variation explained. Hence, dimension 1 was the only strong latent construct underlying the variables.

As evident in **Table 3**, almost all the variables were well represented by the retained dimensions. **For dimension 1**, items such as no ANC (corr=1.62; contr=0.14), home delivery (corr=1.40; contr=0.12), and no health card (corr=1.64, contr=0.12) had strong positive coordinates and contributions, while yes PNC (corr=-1.28, contr=0.04) and two doses of tetanus vaccination of the mother (corr=-1.44, contr=0.08) had strong negative coordinates and contributions. For **dimension 2**, more than average birth weight (corr=-1.18, contr=0.02), complete immunization of the child (corr=-5.46, contr=0.12), and ever breastfeeding (corr=-1.93, contr=0.19) had strong negative coordinates and strongly shaped the dimension.

**Table 3.** Multiple correspondence analysis of the selected eight factors related to maternal and child health indicators.

| Variables | Overall |  |  | Dimension 1 |  |  | Dimension 2 |  |  |
| --- | --- | --- | --- | --- | --- | --- | --- | --- | --- |
|  | Mass | Quality | %inertia | Corr | Sqcorr | Contr | Corr | Sqcorr | Contr |
| **ANC** |  |  |  |  |  |  |  |  |  |
| No ANC | 0.053 | 0.806 | 0.134 | 1.620 | 0.799 | 0.140 | 0.574 | 0.006 | 0.018 |
| < 4 visits | 0.050 | 0.750 | 0.044 | -0.925 | 0.744 | 0.043 | -0.326 | 0.006 | 0.005 |
| ≥ 4 visits | 0.022 | 0.793 | 0.073 | -1.867 | 0.787 | 0.075 | -0.665 | 0.006 | 0.010 |
| **Delivery** |  |  |  |  |  |  |  |  |  |
| Home | 0.062 | 0.827 | 0.114 | 1.400 | 0.827 | 0.123 | 0.162 | 0.001 | 0.002 |
| Facility | 0.063 | 0.827 | 0.114 | -1.400 | 0.827 | 0.123 | -0.162 | 0.001 | 0.002 |
| **BW** |  |  |  |  |  |  |  |  |  |
| > average | 0.018 | 0.950 | 0.013 | -0.921 | 0.860 | 0.015 | -1.181 | 0.090 | 0.025 |
| Average | 0.079 | 0.878 | 0.005 | 0.275 | 0.877 | 0.006 | 0.035 | 0.001 | 0.000 |
| <average | 0.028 | 0.357 | 0.004 | -0.187 | 0.204 | 0.001 | 0.639 | 0.153 | 0.012 |
| **Health card** |  |  |  |  |  |  |  |  |  |
| No | 0.046 | 0.828 | 0.118 | 1.645 | 0.806 | 0.124 | -1.077 | 0.022 | 0.053 |
| Yes | 0.079 | 0.828 | 0.068 | -0.951 | 0.806 | 0.072 | 0.622 | 0.022 | 0.031 |
| **Breastfeed** |  |  |  |  |  |  |  |  |  |
| Ever | 0.051 | 0.731 | 0.015 | 0.216 | 0.119 | 0.002 | -1.936 | 0.612 | 0.191 |
| Never | 0.001 | 0.107 | 0.001 | -0.365 | 0.105 | 0.000 | -0.184 | 0.002 | 0.000 |
| Still | 0.073 | 0.723 | 0.011 | -0.144 | 0.108 | 0.002 | 1.363 | 0.616 | 0.135 |
| **PNC** |  |  |  |  |  |  |  |  |  |
| No | 0.094 | 0.979 | 0.013 | 0.411 | 0.964 | 0.016 | 0.201 | 0.015 | 0.004 |
| Yes | 0.029 | 0.969 | 0.038 | -1.287 | 0.952 | 0.047 | -0.690 | 0.018 | 0.014 |
| DK | 0.002 | 0.373 | 0.003 | -0.770 | 0.368 | 0.001 | 0.360 | 0.005 | 0.000 |
| **Immunize** |  |  |  |  |  |  |  |  |  |
| None | 0.073 | 0.661 | 0.026 | -0.391 | 0.333 | 0.011 | 1.535 | 0.328 | 0.173 |
| Incomplete | 0.048 | 0.736 | 0.036 | 0.704 | 0.503 | 0.024 | -1.893 | 0.232 | 0.170 |
| Complete | 0.004 | 0.674 | 0.015 | -1.148 | 0.275 | 0.005 | -5.465 | 0.399 | 0.124 |
| **Tetanus** |  |  |  |  |  |  |  |  |  |
| No dose | 0.075 | 0.881 | 0.060 | 0.947 | 0.870 | 0.068 | 0.421 | 0.011 | 0.013 |
| <2 doses | 0.008 | 0.647 | 0.019 | -1.394 | 0.640 | 0.016 | -0.577 | 0.007 | 0.003 |
| ≥2 doses | 0.041 | 0.888 | 0.076 | -1.447 | 0.877 | 0.087 | -0.652 | 0.011 | 0.018 |

**Abbreviations:** ANC: Antenatal Check-up; BW: Birth Weight; Corr: Coordinates; Contr: Contribution; DK: Don't Know; Sqcorr: Squared Correlations; PNC: Postnatal Check-up

Antenatal check-ups, place of delivery, possession of health cards, PNC, and tetanus toxoid vaccination were strongly discriminating items for **dimension 1**. Likewise, immunization, breastfeeding, and birth weight were strongly discriminatory for **dimension 2**. Hence, **dimension 1** primarily focused on maternal healthcare utilization, and **dimension 2** focused on child healthcare. Since dimension 1 explained most of the variation, we employed a general linear model that considered only the first dimension. Both the assumptions of normality of the standardized residuals and heteroscedasticity were violated, as the p-values were <0.05 in both tests; however, there was no multicollinearity, as VIFs were <5 for all predictors. The Q-Q plot of the residuals was a bit S-shaped (**Supp Fig 1**).

The increasing value of dimension 1 reflected poor maternal health care utilization, such as no ANC, home delivery, no health card, no PNC, and no tetanus toxoid injection. The age of women was inversely associated with the increasing value of dimension 1 (β (95% CI): **-0.009 (-0.012, -0.005)) (Table 4)**. Women from the higher wealth indices were inversely associated with the increasing value of dimension 1 compared to the poorest wealth index. Likewise, there was a 0.447-unit decrease in dimension 1 value among women with secondary or higher education (p<0.001). Similarly, the use of unimproved fuel and drinking water use was positively associated with the increasing value of dimension 1.

**Table 4.** General Linear Model of Regression to find association between the first dimension (continuous dependent variable) and explanatory variables.

| Explanatory variables | Dimension 1<br>Beta coefficient (95% CI)* | p-value |
| --- | --- | --- |
| **Area of residence** |  |  |
| Rural | 0.023 (-0.018, 0.064) | 0.276 |
| Urban | *Reference* |  |
| **Age of women (years)** | **-0.009 (-0.012, -0.005)** | **<0.001** |
| **Sex of the youngest child** |  |  |
| Girl | **0.049 (0.023, 0.076)** | **<0.001** |
| Boy | *Reference* |  |
| **Wealth index** |  |  |
| Poorer | **0.049 (0.007, 0.091)** | **0.02** |
| Middle | **-0.102 (-0.146, -0.058)** | **<0.001** |
| Richer | **-0.250 (-0.297, -0.203)** | **<0.001** |
| Richest | **-0.257 (-0.324, -0.191)** | **<0.001** |
| Poorest | *Reference* |  |
| **Education status of women** |  |  |
| Primary | **-0.350 (-0.399, -0.301)** | **<0.001** |
| Secondary and higher | **-0.447 (-0.491, -0.402)** | **<0.001** |
| Illiterate | *Reference* |  |
| **Type of toilet facility** |  |  |
| Pit Latrine | **0.114 (0.071, 0.158)** | **<0.001** |
| Open defecation | **0.355 (0.297, 0.413)** | **<0.001** |
| Flush Latrine | *Reference* |  |
| **Drinking water source** |  |  |
| Well or river (unimproved) | **0.219 (0.191, 0.247)** | **<0.001** |
| Piped or tap (improved) | *Reference* |  |
| **Type of cooking fuel** |  |  |
| Solid (unimproved) | **0.211 (0.172, 0.251)** | **<0.001** |
| Gaseous (improved) | *Reference* |  |
| **Number of children** | **0.045 (0.037, 0.054)** | **<0.001** |

\*A $p$ -value<0.05 was considered a statistically significant value; all the significant associations were highlighted in bold. AIC: 2.68; BIC=-171456.3; Log pseudolikelihood=-25568.749; deviance=16314.78; residual deviance=0.85
**Abbreviations:** CI: Confidence Interval

## Discussion

The present study found that most women were uneducated or educated only up to the primary level, both in rural and urban areas. The utilization of maternal health services was worse in rural areas and poor in urban areas, too. Women had limited access to antenatal and postnatal services and institutional delivery, with most children completely unimmunized or incompletely immunized. Poor socio-demographic characteristics were positively associated with poor maternal and child health outcomes.

With the poorest maternal and child health indicators in South-Central Asia, Afghanistan is a country that needs urgent actions and support from international agencies to prevent a huge toll of maternal and child deaths in the coming years. Most women were illiterate and had an average of 4 children by the age of 30 years. This could be understood from the phenomenon of widely prevalent early marriage in Afghanistan, which is a part of their culture, and largely due to poverty, illiteracy, women’s disempowerment, poor socio-economic situation, and pressure from parents and society [17]. The use of unimproved toilet facilities (pit toilets) and open defecation accounted for nearly 90% in rural areas and 60% in urban areas. Likewise, nearly 60% of women were drinking water from an unimproved source. It is not uncommon to find increased use of unimproved toilets and drinking water facilities in countries of South Asia and Africa [17, 18].

The uptake of the recommended 4 ANC services in Afghanistan is only 13% in rural and 28% in urban areas. Our finding concurs with other studies from Afghanistan, which have also reported that only half of these ANC visits were conducted by skilled birth attendants [19, 20]. Access to regular ANC visits is a step toward ensuring safe motherhood, helping diagnose and treat any pregnancy-related complications, promoting institutional delivery, and providing preventive health education and promotion services [19]. Recently, the World Health Organization (WHO) recommended a minimum of 8 ANC visits to support a positive pregnancy experience and early detection of high-risk pregnancies [21]. The present study findings highlight a huge gap in the opportunity for the health system to assess high-risk pregnancies and complications among women. Although the youngest child was 2 months old, only half of the women (∼55%) were still breastfeeding their children at the time of the survey. This is supported by the evidence from previous literature reporting the influence of cultural beliefs and practices, misconceptions, and social norms that promote pre-lacteal feeding, late initiation of breastfeeding, and colostrum avoidance in Afghanistan [22].

Contrary to the previous studies reporting nearly 46% full immunization rate among children, we found that only 7% of children were fully immunized against 5 vaccines (BCG, DPT-1, 2, 3, and Measles) [23–25]. Plausible reasons for the difference could include the lack of data on pentavalent vaccination in our dataset. In addition, the criteria used for full immunization by us included vaccination against all 13 vaccines contrary to other studies. Furthermore, variations in the place of delivery, maternal age, parental education, socioeconomic status, and media exposure may further contribute to the differences observed between studies [26]. Full immunization of children is a critical component of UHC and needs to be attained for not just reducing mortality from vaccine-preventable diseases, but also to protect children against malnutrition [25]. Nearly two-thirds of women had a health card, which helped check the vaccination status of children. The DHS survey report of 2015 highlighted that mothers who had a boy child, lived in urban areas, were educated, and belonged to a wealthy quintile had a higher prevalence of carrying a health card [24].

Likewise, mothers who were younger, had a girl child, belonged to the lower wealth quintile, and were illiterate, had a higher probability of dimension 1 that reflected poor maternal and child health services utilization (p<0.05). Furthermore, women who did not have access to improved drinking water and toilet facilities, and improved fuel source had a higher risk of dimension 1 (p<0.05). Empirical evidence suggests the inevitable role of these socio-demographic characteristics in determining the demand for maternal and child health services [19, 20]. These factors form the first and most critical step of the ‘three-delay model,’ i.e., delay in seeking care, in accessing health services [27]. Furthermore, Christou *et al*., 2020, supported the low uptake of maternal health services by highlighting the poor health-seeking behavior of Afghan women, despite their availability [28]. Lack of financial resources, long distances to facilities, and limited transport have also been highlighted by Azimi *et al*., 2015 [29].

Although most health and socio-demographic indicators performed worse in rural areas than in urban areas in the present study, the difference was not statistically significant. However, other studies highlighted geographical inequities in maternal healthcare utilization in Afghanistan [20]. This is largely explained by the lack of availability of healthcare facilities, access to educational institutions, public awareness, transportation infrastructure, and access to the internet in rural areas [30]. Needless to mention these poor socio-demographic determinants culminate in high fertility rates, maternal and child mortality and morbidity, and low life expectancy in many countries of Africa and Asia [31].

Afghanistan made significant strides in many maternal health outcomes after the uprooting of the Taliban government in 2001, including increases in the institutional delivery rate by 17% and deliveries by skilled birth attendants by 19% from 2001 to 2015; however, the progress was reported to be uneven [32]. And the comeback of the Taliban government in 2021 has once again jeopardized the progress made by the country in two decades. This demands a strong determination by international agencies to negotiate with the local Taliban government and health professionals, and reinstate the basic package of maternal and child health services in the country.

### Strengths and limitations

The present study assessed the DHS data of maternal and child healthcare service utilization in Afghanistan, given that there is limited literature but the absolute dire need for it in this domain. The key parameters that primarily define maternal and child healthcare utilization were selected, and MCA was applied to construct a latent variable that reflects all selected indicators. However, the study had multiple limitations. The data for the present study were collected while the country was engaged in an ongoing conflict. As a result, the DHS could not capture data from the remotest areas of the country. The findings may not be generalized to areas outside Afghanistan due to the unique socio-cultural dynamics, norms, and traditions of the country.

## Conclusions

The present study concluded that most women were uneducated in both rural and urban areas in Afghanistan. Most maternal and child health indicators were poorer in rural areas than in urban areas, though this difference was not statistically significant. Women who were younger, had a girl child, belonged to the lower wealth quintile, and were illiterate, and did not have access to improved drinking water and toilet facilities, and improved fuel had a higher probability of poor maternal and child health indicators.

## Declarations

### Availability of Data

The anonymized dataset is available on USAID DHS webpage (https://dhsprogram.com/). The authors are not allowed to share the data with others.

### Author’s contribution

KM and NS designed the study. KM and SS carried out the statistical analyses. SS wrote the first draft. All authors (KM, NS, and SS) contributed with intellectual content, revised, and approved the final version of the manuscript before submission.

### Copyrighted figures

The authors declare that they have not used any copyrighted figures.

### Dual publication

No element in the manuscript have been published or are under consideration for publication elsewhere.

## Acknowledgements

We would like to thank the study participants who provided their information during the survey and USAID for providing us the data for analysis.

